# Body Shaming vs. Body Positivity: Exploring What Words Hurt and What Words Heal

**DOI:** 10.1101/2023.05.09.23289725

**Authors:** Valerie Wansink, Brian Wansink

**Affiliations:** Lansing Central School District; Retired, Cornell University

## Abstract

What can a health professional say to a bullied and body shamed child? This research elicited and categorized the words that were reported as being most memorably hurtful and helpful to 341 young people (79.5% female; average age 25.03 years) who had been body shamed. The most memorably hurtful comments generally involved either vivid comparisons or derogatory nicknames. Fortunately, the supportive words of health professionals, peers, and parents can heal. They include 1) *reframing comments* that redirect attention to positive features (such as feminine, healthy, etc.), 2) *impact-related comments* that emphasize how a physical feature influences or is admired by other people (such one’s eyes or smile that “lights up the room”), and 3) *identity-related comments* that redefine one’s physical identify or self-concept (such as striking or mesmerizing). Knowing the type of words that were helpful in healing can be useful to healthcare professionals as well as parents and peers.

---

Body shaming, fat shaming, and weight bullying can have a dramatic impact on a person’s self-esteem (1,2). A key question in this exploratory research is what are types of name calling or verbal bullying are most likely to have a long-lasting negative impact on a young person (3), and what types of comments can be made by a supporter – a health professional, parent, or peer – to help counterbalance or temper this impact? In short, what words hurt and what words heal.

Body shaming can cause lasting psychologically damage (4). Although this can happen at all ages (5), it may be most pronounced and harmful when a person is young (6,7). Body shaming and bullying is correlated with low self-esteem, poor school performance, absenteeism, self-handicapping, and even suicide (4,8), and being bullied as a child is a powerful predictor of exaggerated weight gain in some people (9). Shaming and bullying is no longer just a problem in middle and high school (10) because it now extends to the internet and to social networking sites (11). It has a 24/7 impact.

Research has not yet codified the actual types of words that are most memorably painful to a person (12), nor have they codified the types of words that have been most helpful to their recovery. Knowing this could inform potential supporters – health professionals, parents, or peers – on how to help a bullied person better counter (13) and cope (14) with body shaming. Moreover, these findings can be useful to the peer or parent who is asking themselves “What do I say?” or “How can I help?”

To help answer these questions, a survey targeted at body shamed individuals asked them to explain what was said to them that was most hurtful and what did supporters say to them that was most helpful in their healing process. After analyzing these open-ended answers, the results are discussed with an eye toward how they might be used to guide health professionals to help a young person who has been body shamed or fat bullied.

## Background

Words hurt, but it is not clear what words hurt the most (15). Whereas experimental research shows that even short-term teasing can negatively alter one’s mood (16), more intense and malicious verbal bullying could last for many years (17). It can lead to shame (18), stress (19), eating disorders (20), obesity (9), binge eating (21), and depression (22). This can affect all genders and is believed to be both more prevalent and more serious than in the past (1) due to the large prevalence of 24/7 social media (23).

Being bullied can negatively change one’s self-concept (15). There are a number of key dimensions of a person’s self-concept (5) and there are several ways adolescents define themselves and their identity (18, 24). For instance, they might define themselves as a debater, a clarinet player, a diligent student, or as a loyal friend. Alternatively, if a person has been ridiculed by their weight or height, they may come to define themselves mostly by this physical trait, and not by positive descriptions of themselves, such as their social skills, or other talents (25–29). Such “self-talk” could also tend to negatively influence life decisions they make.

This exploratory research investigates these questions, starting with “What types of words hurt the most?” In looking toward gender-inclusive solutions, we also determine what type of words could be useful to those supporters who want to help. That is, we examine what types of words <u>help</u> the most?

## Method

An online omnibus survey was targeted at recent high school graduates (18 years and older). They were recruited through a news release that described the purpose of the study and which was sent to youth health and lifestyle bloggers. A survey link was provided to bloggers and to others who circulated this link through social media and through email lists. These bloggers and list managers then promoted the topic and the link in ways that were intended to attract people who were familiar with body shaming.

Upon arriving to the survey’s website, visitors were told “You are invited to participate in a research study that seeks to understand the issue of body shaming (the action or practice of humiliating someone by mocking or making critical comments about their physical appearance) among young adults and how to preempt, cope, and stop it.” Those who indicated they were 18 years or older and who signed their consent were then allowed to begin the survey. The survey and study had Institutional Review Board approval from Cornell University.

The questions generally referred to body shaming that occurred in adolescence (middle school or high school). Of those people who completed most or all of the survey, 61.4% were between 18-22. This was the targeted age range because it has been the general age-range in cyber bullying and teasing research (30). It was believed that their initial experiences with body shaming would be recent enough to be remembered, but distant enough to allow for reflection (31).

They were asked open-ended questions about their specific experiences with body shaming, including the hurtful comments that were made as well as the helpful comments given by supporters. They were also asked exploratory close-ended questions, such as their familiarity with the term “body shaming,” and they were asked to indicate what level of body shaming they had experienced as an adolescent.

Of the 341 people who started the survey, 166 (48.7%) completed all or most of the survey. The median time spent on the survey was around 10 minutes, and those who provided the most in-depth open-ended responses took the most time with the survey. Of those completing the survey, 79.5% identified as females, one as transgender, and the rest as males. The average age of those completing the survey was 25.03 years (ranging from 18-62).

Based on the responses of all genders and ages, we developed an inclusive coding system so the comments of the respondents could be content analyzed and then categorized into discrete categories. A Q-sort procedure was used to determine the coding categories for each open-ended question. After determining these coding categories, two coders in the age range of interest coded the open-ended responses. Any resulting disagreement between the two coders final responses was then discussed and resolved.

## Results

Inclusive coding categories were developed for both hurtful and helpful comments. Categories of hurtful comments included vivid comparisons and derogatory nicknames. In contrast, categories of helpful comments included reframing comments, impact-related comments, and identity-related comments. After first examining the physical features targeted by attackers and supporters, we will then focus on the types of comments that were most memorably hurtful and helpful.

### The Physical Features Most Mentioned by Attackers and by Supporters

Of initial interest are the specific physical features mentioned by attackers and those specific physical features mentioned by supporters. As Table 1 indicates, most of hurtful body shaming comments were targeted at being fat (25.2%), short (8.5%), or skinny (7.1%). In total, nearly a third (32.3%) of the most hurtful comments specifically focused on a person’s weight, and four out of five (78.0%) were about being overweight.

**Table 1:**
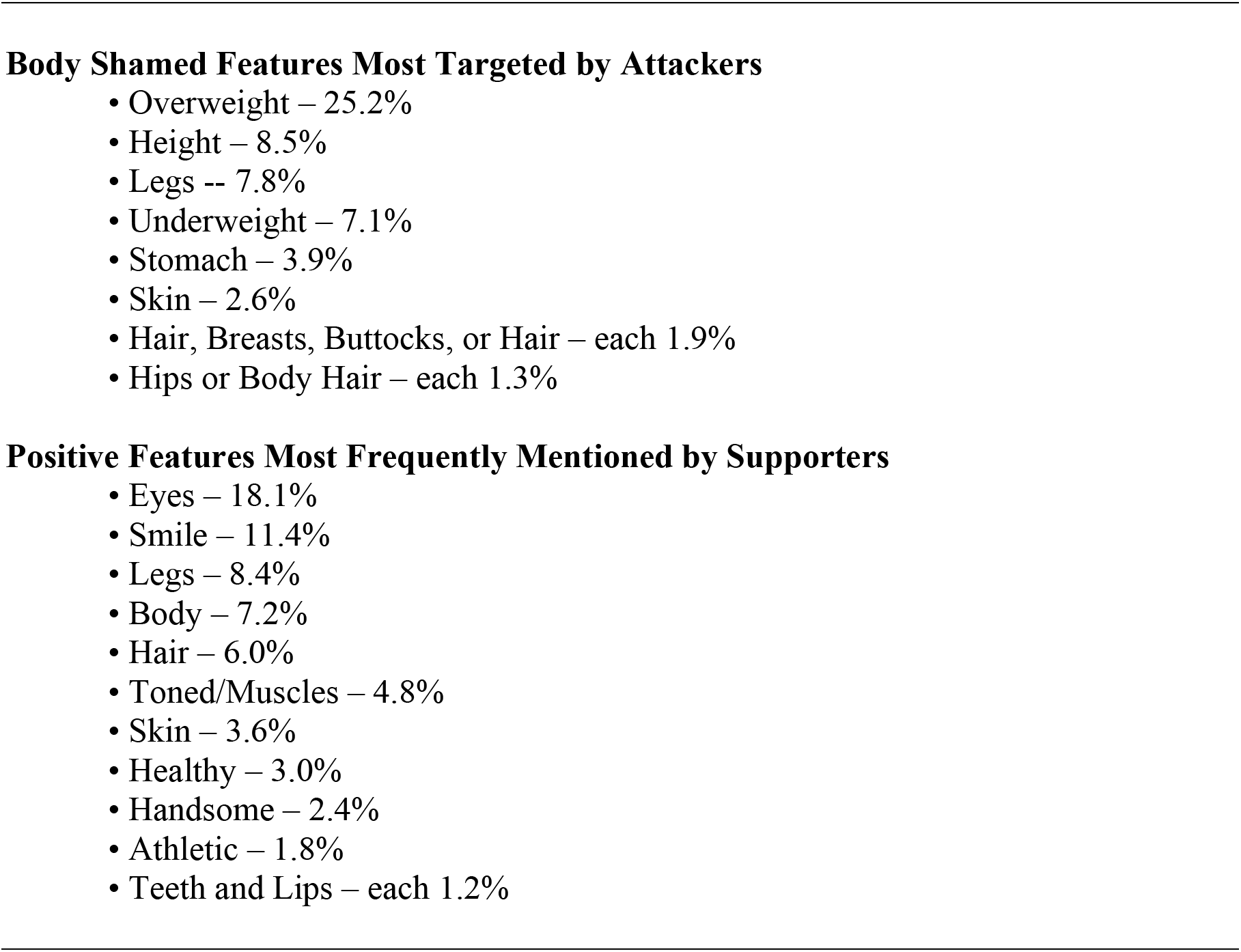
The Physical Features Most Mentioned by Attackers and by Supporters.

Interestingly, people who had been body shamed also vividly remembered the positive words that supporters had mentioned – in some cases nearly 50 years ago. The most common category focused on positive physical features *unrelated* to those that were bullied – such as eyes (18.1%) or smile (11.4%), which (along with teeth and lips) comprised nearly a third (31.9%) of all the most positively remembered physical features.

### Hurtful Negative Comments Include Exaggerated Comparisons

One category of hurtful comments that stood out were those that made an exaggerated comparison or analogy about the specific physical trait it targeted. These included comparing a person’s size to that of a whale, comparing one’s thighs to trees, or one’s stomach to that of a pregnant woman. Although these comments about being overweight comprised 25.2% of all comments, 7.1% were shamed and bullied for being underweight, such as being compared to a skeleton or as having “chicken skinny” legs (see Tables 1 and 2). For instance, one girl remembered people describing her like a board, saying “They said they can’t tell the difference between my front and back because I am quite skinny and have few curves.”

**Table 2:**
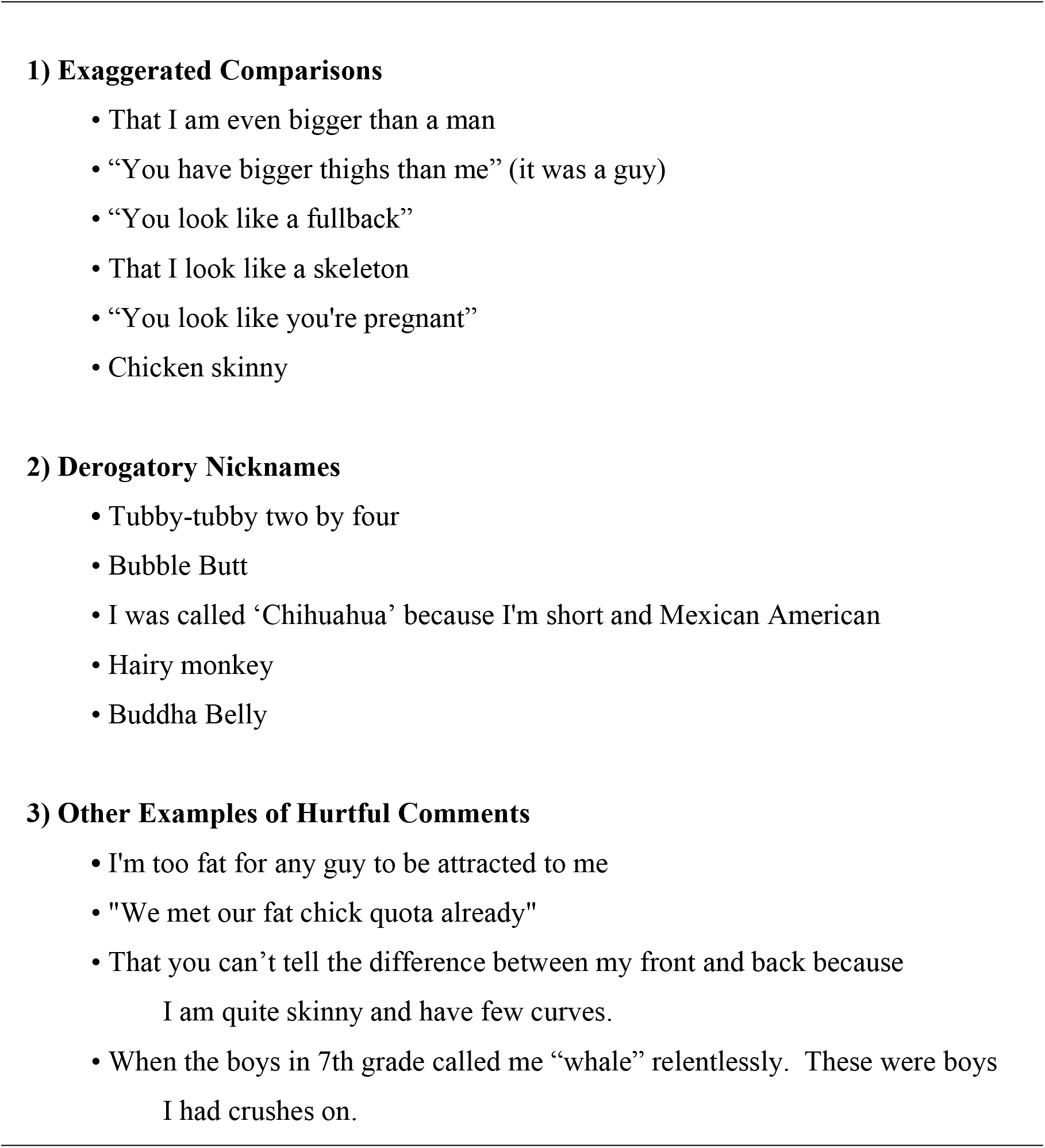
Categories and Illustrations of Memorable Negative Comments.

A second common category of body shaming that was commonly mentioned included derogatory nicknames (see Table 2). This category frequently included alliterative names like Buddha Belly or references to popular culture (Hairy Potter). One person recalled being called “Chihuahua” because they were short and Mexican American.

Hurtful comments were sometimes specified as coming from a relative or from an admired person. These included a father insensitively joking about a girl’s hair, a mother commenting on breasts, or a brother teasing about weight. One girl hurtfully recalled a time “When the boys in 7th grade called me “whale” relentlessly. These were boys I had crushes on.”

### Helpful Positive Comments Include Reframing and Redirecting

One type of memorable or helpful comment were *reframing comments*, which reframed the physical feature that had been attacked. It did not deny what was said but instead reframed it by focusing on one’s “hourglass” figure, one’s muscle tone, or as one being “healthy” or “athletic.” In addition to reframing, other memorable comments specifically redirected one’s focus onto physical features that were *unrelated* to the features that was shamed. As Table 1 indicated, the most frequently mentioned examples of this were a person’s eyes and smile.

A second category of helpful comments were *impact-related comments* that emphasized how a person’s features impact other people in a positive way. These included comments such as how a person’s smile, eyes, or laugh was “contagious,” “infectious,” “lit up a room,” or “radiated joy.” It also included comments that showed that they had traits that other people envied or wished they had (such as “I wish I had your eyes”). For examples, being told you have a wonderful smile might be memorable partly because of who said it. One person said, “My husband always describes me to others as the pretty girl with the big smile.” Another person simply said, “My father complimented my smile.” These were very simple but helpful comments coming from an important person.

A third category of memorable comment were *identity-related comments* that generally centered on a single, evocative word that reinforced their an overlooked part of their identity more than a specific physical feature. That is, they were “mesmerizing,” “striking,” “exotic,” or “feminine.” Even though these were often stated in conjunction with other words (“mesmerizing beauty”), they were seen by a person as a more general part of a positive identity. Whereas the adjectives in the negative body shaming comments were general and not creative (such as “really fat” or “very short”), the positively remembered comments were more vivid, creative, or artfully used. Some became a tent pole that reinforced a new, positive identity.

## Discussion

As an exploratory study, this usefully specifies and classifies the types of hurtful phrases used in body shaming and bullying, but it also uniquely classifies and provides examples of helpful and memorable phrases. By doing so, it provides a foundation to motivate new research in these areas of hurtful remarks and helpful remarks.

Words hurt, and these preliminary findings show that some types of words leave a more vivid and lasting pain. Follow-up discussions indicated it was not only a general label that hurt (fat, short, skinny, and so on) but it was also derogatory nicknames (Thunder Thighs) and vivid comparisons (whale and Buddha Belly) that were especially painful, possibly because they are very personal, very specific, and they can be easily triggered in a wide variety of ways. That is, the mention of a Harry Potter movie can trigger “Hairy Potter” to mind, or the mention of “whale watching” can trigger a memory of being called a whale and hearing someone make whale imitations during school lunch.

An encouraging insight from these findings is they also suggest what a health professional, parent, or peer might say to help counter the body shaming messages. Table 3 usefully presents the types of positive phrases that have tended to stick with people over the years. One strategy can be to *reframe negative words* (see Appendix) to focus on new or unrelated physical features. A second strategy can be to make *impact-related comments* that emphasize how a physical feature influences others (such one’s eyes or smile that “lights up the room”). A third strategy can be to use *identity-related comments* that can positively redefine one’s physical identify or self-concept (such as being striking or mesmerizing).

**Table 2.**
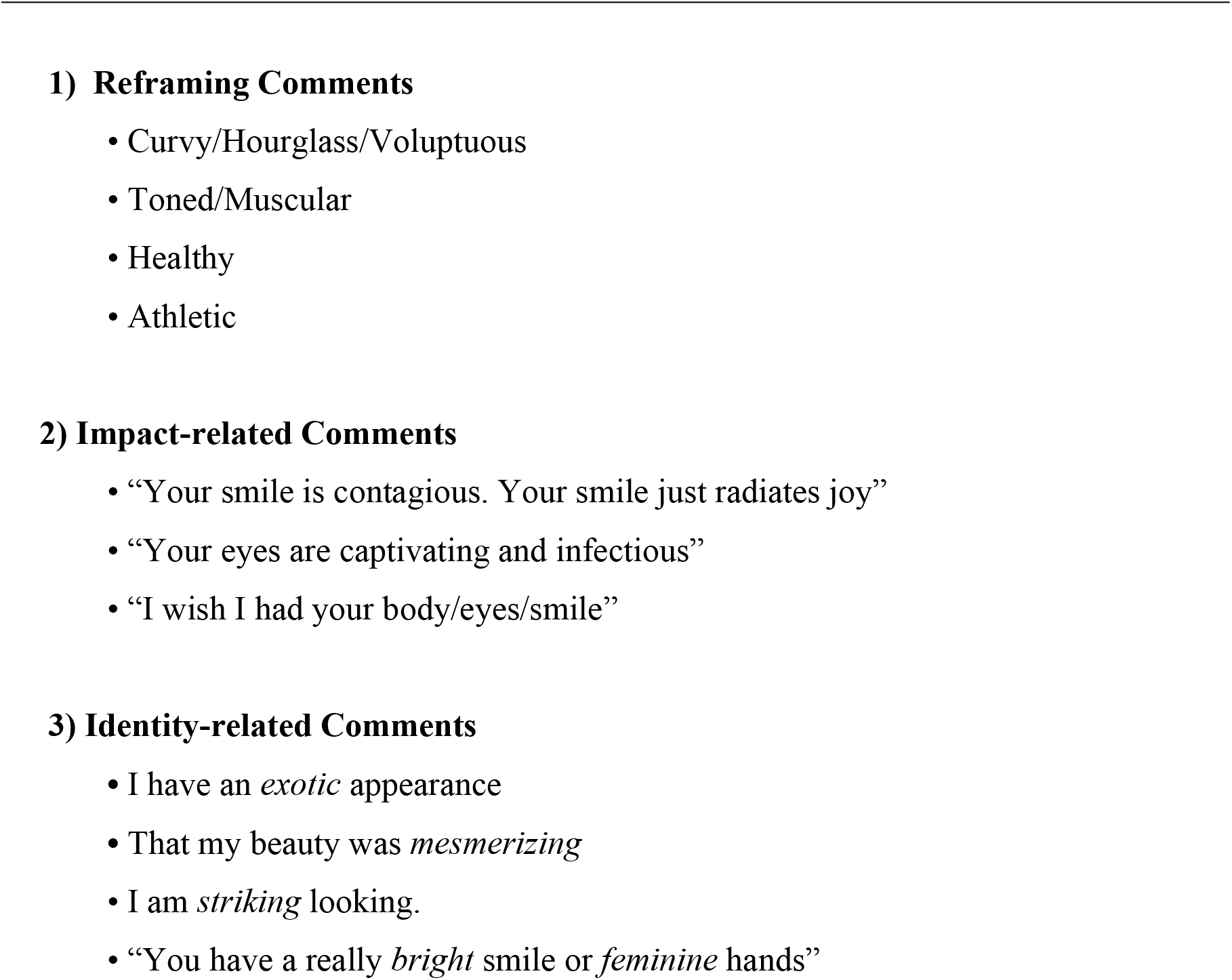
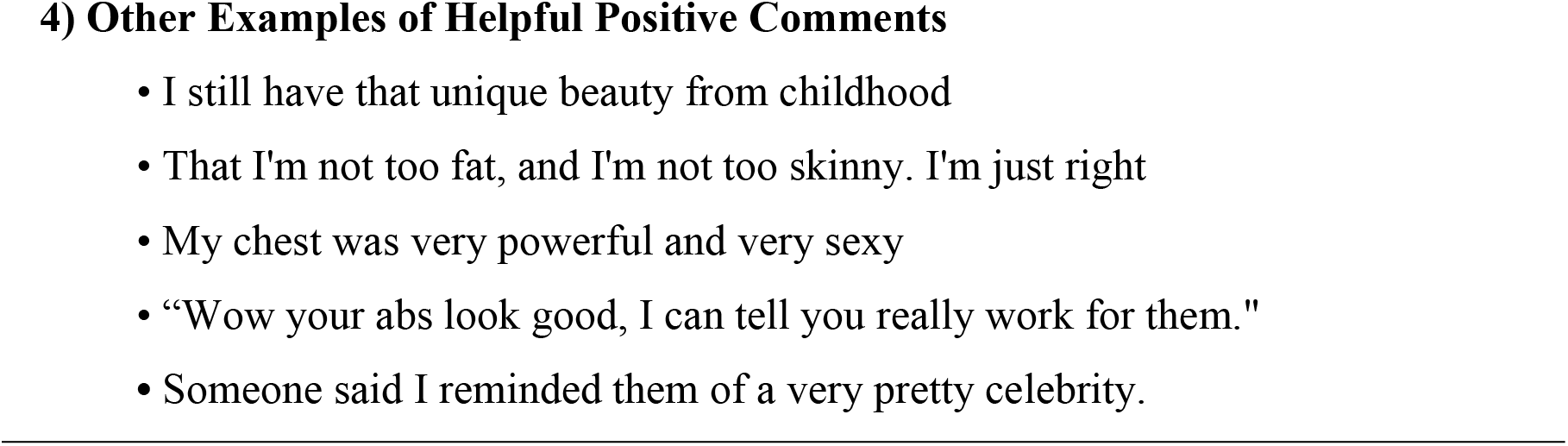
Categories and Illustrations of Memorable Positive Comments.

## Implications

Body shaming can be fought on three fronts: 1) By the individual being attacked, 2) by supporters (health professionals, parents, and peers), and 3) by the institution in which the shaming occurs. First, for an individual who may be vulnerable to body shaming, knowing that there are some predictable categories of comments that are universally hurtful could be reassuring. It shows that the person is not isolated in the way they are attacked and what is said. Knowing that there are many people who have gone through the same type of shaming and who have recovered could make a person feel less alone and more hopeful.

Second, for supporters – health professionals, parents, and peers – it should be helpful to know what specific types of helpful words and what categories of comments might be most memorable and useful to the recovery of their loved one. This is important for anyone who has ever thought “What can I say?” Knowing these categories of helpful words, can give them confidence in stimulating what might be most helpful to the specific case of their loved one. They are not helpless and what they say can have a lasting impression (32).

Interestingly, the types of comments body shamed people remembered as most helpful to their recovery can be can be useful for help professionals, parents, and peers as their prepare for an empowering discussion with a body shamed person. The Appendix is a worksheet titled “Words that that Help Heal Body Shaming.” Supporters can use it to think more carefully about which of the three approaches and which particular words could be useful when preparing for a conversation with a patient or a loved one.

Third, because schools are one common area where in-person body shaming can occur, they can also be a context where these body shaming discussions could occur. Counselors, nurses, teachers, and lunch staff, could be given easy-to-remember guidance on the types of words (comparisons and derogatory names) that are hurtful and the types of words (and strategies) that can be memorable or helpful in recovery (see Appendix). Most easily, giving these front-line workers a basic framework – such as “Recognize and Reframe” – could be useful in stopping body shaming and in helping comfort those who are hurt. In a bigger way, lunchrooms themselves can be altered in ways that help students eat healthier (33–36).

## Limitations and Future Research

This exploratory study usefully highlights several topics that have been missing from earlier literature. Most of those who took this survey were college-aged people who were body shamed during adolescence. Given their closeness to this emotional issue, they could have had the tendency to either exaggerate or underplay (repress) what really happened.

Additionally, this is not a random sample. This population represents a well-educated group sample compared to the larger pool of those who are body shamed. Being well-educated might moderate some of the identity problems associated with being body shaming if it enables a person to build upon a professional identity. That is, success in one area (such as in a profession) could help ameliorate one’s feelings of shortcoming because they were body shamed.

This research shows the importance of capturing the experiences of body shamed people to better understand what might help to heal body shaming: how they dealt with the shamers, what self-talk worked best for them, and how they were able to heal and move on. There is a great deal of power in analyzing their open-ended answers with an eye toward providing specific roadmaps and tools to help others in moving forward (37).

## Conclusion

Body shaming, weight shaming, and bullying can scar a person for life. These results give insights for immediate action. These results show supporter – health professionals, parents, and peers – the ways they can *reframe negative words* to focus on new or unrelated physical features. They show them how to make *impact-related comments* that emphasize how a physical feature influences others (such one’s eyes or smile that “lights up the room”). They also show how *identity-related comments* can be used to positively redefine one’s physical identify or self-concept (such as striking or mesmerizing). With an increase in childhood obesity, body shaming may become even more severe. Hopefully the words here and that can be generated from the worksheet in the Appendix can help.

## Data Availability

All data will be made available on line and with the publisher upon publication of the manuscript

## APPENDIX

### Words that that Help Heal Body Shaming Worksheet

People who have recovered from being body shamed often point to helpful comments people made to them. This worksheet helps you identify which of three different approaches in offering helpful words might be most useful to someone who has been body shamed.

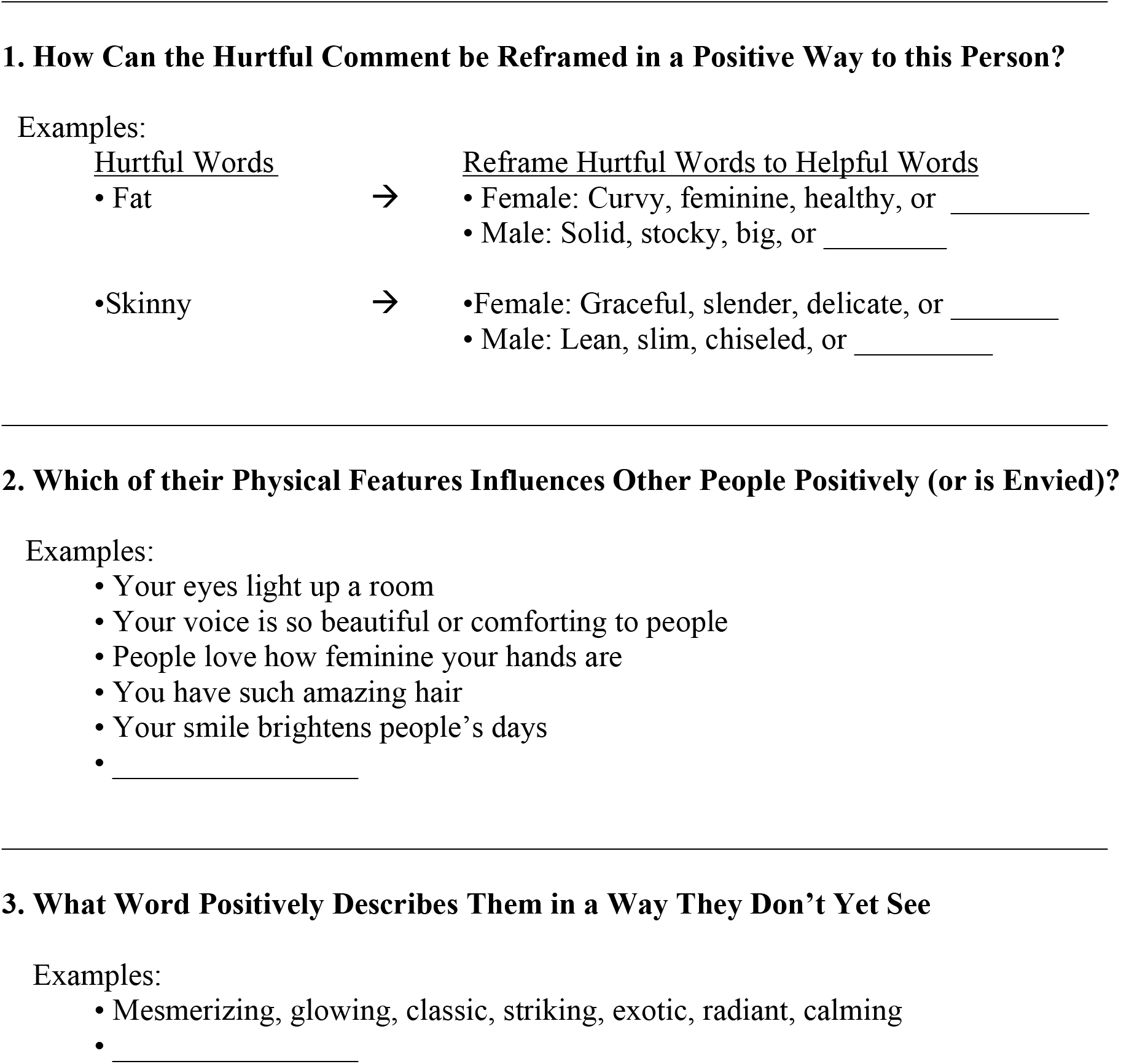

